# Schizophrenia, inflammation and temporal change in brain morphology: an omnigenic Mendelian randomization study

**DOI:** 10.1101/2023.01.17.23284695

**Authors:** Hongyan Ren, Yunjia Liu, Yamin Zhang, Qiang Wang, Wei Deng, Xiaohong Ma, Liansheng Zhao, Xiaojing Li, Pak Sham, Tao Li

## Abstract

The last decades of research in schizophrenia witnessed a shift of etiological speculation from neurotransmitters to inflammation. However, identifying definite inflammatory effectors of schizophrenia remains elusive due to confounding factors such as medication and metabolic status. To tackle this issue, we carried out omnigenic-based Mendelian randomization (MR) analysis to explore the inflammatory responses of schizophrenia and the brain morphological consequences caused by these SCZ-triggering inflammation responses. Our results identified seven SCZ-triggering inflammation markers, with P values surviving the Bonferroni multiple comparisons (B_NGF, P = 1.45 × 10^−8^; GROA (CXCL1) P = 1.15 × 10^−4^; IL8, P = 3.64 × 10^−7^; MCSF, P = 9.30 × 10^−4^; MCP3 (CCL7), P = 1.3 × 10^−6^; TNF_β, P = 3.63 × 10^−4^; CRP, P = 1.71 × 10^−32^). Further, three of them, GROA (CXCL1), IL8 and CRP, could lead to significant linear change rate of brain morphologies, especially white matter in both cerebral and cerebellum. Our study is the first to use an omnigenic conceptual framework to capture the immune pathology of schizophrenia. Although future studies adopting a different methodology are needed to validate our results, our study provides another piece of evidence that extensive and low-grade neuroinflammation exists in schizophrenia and that some of these inflammation markers could be potential targets for the precise diagnosis and treatment of schizophrenia.

## 1. Introduction

Schizophrenia (SCZ) is a complex disorder with a genetic architecture involving both polygenicity and pleiotropy [1, 2]. The rapid advance in genotyping and sequencing technology, combined with multinational collaborations in collecting samples, epitomized by Psychiatric Genomic Consortium (PGC), facilitates the genome-wide scanning of risk variants for SCZ in a well-powered sample. The arising new results shed new light on the architecture and pathogenesis of SCZ: Rather than involving hundreds or thousands of genes, the seminal paper by Boyle et al., for the first time, proposed an omnigenic model, i.e. in the cell types that are relevant to a disease, essentially all genes contribute to the condition [3]. Meanwhile, three waves of genome-wide association studies (GWASs) by PGC hint at a critical role inflammation/infection might play in the pathophysiological mechanism of SCZ, with the most associated genomic regions pointed to the MHC region on the chromosome 6 [2, 4, 5].

Despite the multi-lined evidence showing the involvement of neuroinflammation in SCZ [6, 7], identifying the inflammatory markers indicative of SCZ remains elusive. The most detectable and widely distributed inflammatory molecules in the peripheral blood system are cytokines and chemokines, both chemical mediators that regulate immune cell homeostasis, and coordinate signal-dependent immune responses [8]. The last two decades witnessed a conceptual shift in framing the relationship between the brain and immune system. Not only is the brain not off-limit from the immune system, the cells in the brain, like neurons and microglia, release the molecules like cytokine in both maintaining normal function and diseases [9]. In schizophrenia, researchers have already found evidence that cytokines, such as C-reactive protein (CRP) IL-1β, IL-6, IL-12, interferon-γ, and tumour necrosis factor α (TNF-α) etc., were associated with endophenotypes and different disease status of schizophrenia [10, 11].

Although these results are promising, they are not always consistent due to the heterogeneity of methodology and moderate study sample size. At the same time, many factors could confound the investigation of cytokine levels in the peripheral blood in schizophrenia, such as antipsychotics, metabolites, the co-morbid inflammatory condition and age [12, 13]. Besides, the statistical method most often employed in association studies is generalized linear regression; there have been heated debates regarding causal inference of regression analysis. Linear regression implies causality only if the covariates are from a controlled experiment and the experiment isolates the hypothesized causal factor well [14]. Unfortunately, most association studies with cytokine levels as covariates were conducted in a non-experimental controlled manner. Against such a reality, Mendelian randomization (MR) provides a valuable tool for detecting a causal relationship between phenotype pairs, generating a more robust causal estimate due to the immutability of genotypes as instrument variables (IVs, randomization at birth) [15]. In recent years, two-sample MR, which leverages the publicly accessible summery-level data from GWAS, gains more popularity due to free access to a vast resource of GWAS results of various phenotypes or traits [16]. Unlike one-sample MR, two-sample MR does not need the effect of the IV-exposure association and the IV-outcome factor association to be collected in the same sample of participants, which could evade the confoundings such as “winner’s curse” [17]. Hartwig et al. adopted such an approach to draw a quantitative causal link between CRP (C-reactive protein), IL1 (interleukin 1) and the lifetime risk of schizophrenia [18]. Although their results are of great potential with a rigorous methodology, there are a few limitations the authors have yet to address. Firstly, Hartwig et al. used the genetic variants passing the genome-wide significance threshold (5 × 10^−8^) as IVs, which could only explain a small proportion of the phenotypic variance of complex traits, thereby predisposing the study toward wear instrument bias [19]. Secondly, the study only focused on a few candidate cytokines. Gradually accumulating evidence suggests that not one or two cytokines but cytokine networks might be involved in the pathophysiology of schizophrenia [20].

Therefore, the current study takes the genetic architecture of SCZ into account and uses an omnigenic model to answer the following two questions: (1) What is the immune response caused by the pathogenesis of schizophrenia in terms of inflammatory molecules, including cytokines, chemokines and CRP (SCZ-triggering inflammatory markers)? A recent study by Enhancing NeuroImaging Genetics through Meta-Analysis (ENIGMA) consortium shed light on the genetic variants associated with the longitudinal changes in brain structure [21]. Further, since various studies found systemic inflammation as a predictor of brain aging [22], we also try to answer the second question (2) What is the effect of the SCZ-triggering inflammatory markers on the longitudinal brain changes? (Fig 1).

**Fig. 1:**
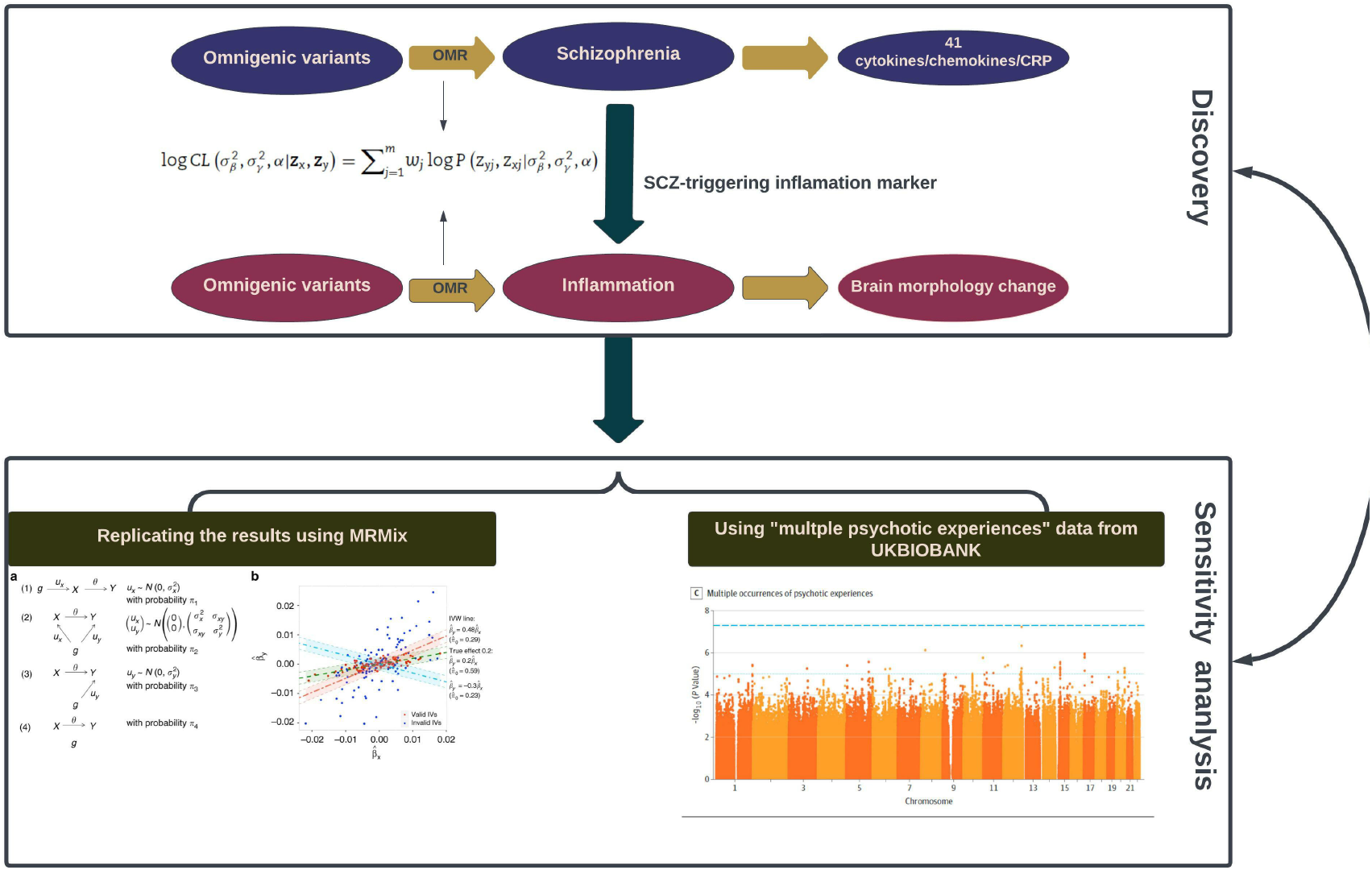
Schematic diagram of analysis framework in our current study

## 2. Materials and methods

### 2.1. Data source

For the MR analysis of SCZ ⇒ inflammation, our study used the summary statistics of schizophrenia GWAS conducted by Trubetskoy et al. involving 76,755 cases and 243,649 controls of European ancestry as the exposure (SCZ) [5]. For the inflammation outcomes, we used the summary statistics of GWAS of 41 cytokines/chemokines in the peripheral blood of 8293 healthy volunteers of European ancestry [23]. As for the CRP, we chose the summary statistics of the latest CRP GWAS conducted on 427,367 participants of European descent [24]. No notable sample overlapping was found within three datasets (SCZ, cytokines/chemokines and CRP).

The SCZ-triggering inflammatory markers identified from the analysis described above were taken to the second round of MR analysis of inflammation ⇒ longitudinal morphology change in fifteen brain regions (amygdala, caudate, cerebellum gray matter, cerebellum white matter, cerebral white matter, cortical gray matter, hippocampus, lateral ventricles, cortical mean thickness, nucleus accumbens, pallidum, putamen, surface area, thalamus, total brain volume). For the outcome measure, we used the GWAS summary statistics of longitudinal changes in brain structure carried out by the ENIGMA consortium in a sample of 15,640 individuals of European descent [21]. To our knowledge, there was no sample overlapping between these two datasets.

### 2.2. Two-sample MR analysis

Due to the easy access to the GWAS results, many methods have been developed to conduct the two-sample MR analysis. However, the majority of these methods still require (1) using SNPs with significant association at the genome-wide level as IVs, and (2) the selected IVs should be independent of each other, i.e. linkage equilibrium (LD). However, as mentioned earlier, schizophrenia, like many other complex diseases/traits, is a polygenic, even omnigenic disease. Further, the independent IVs in a less perfect LD with the true causal ones could lead to power loss. To tackle the issues hereof and increase the power of causal inference, Lu et al., based on the concept of the omnigenic model, developed a new analysis framework for the two-sample MR studies, the omnigenic Mendelian randomization (OMR) [25]. Specifically, OMR relaxes the relevance and independence assumptions by (1) including all genome-wide SNPs to serve as IVs. and (2) by explicitly including an additional term, *G*_*y*_γ, in the equation below to control for the potential horizontal pleiotropy.

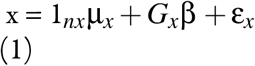

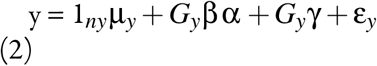

In the equation above, 1 represents a vector of 1’s with the corresponding dimensionality shown in the subscript, µ_*x*_ is the intercept for the exposure model; µ_*y*_ is the intercept for the outcome model; nx and ny are the sample size of GWASs for exposure measure and outcome measure, respectively; β is the genotype effects on the exposure variable; γ is the horizontal pleiotropy effects of SNPs on the outcome variable; α is a scalar that represents the causal effect of the exposure variable on the outcome variable; x and y are residual error vectors of size nx and ny, respectively. The OMR constructs a log composite likelihood as a weighted combination of the log marginal likelihood. Both simulation and real dataset analysis showed that compared with other methods, OMR showed superiority over other methods in controlling type I error and horizontal pleiotropy effects across different genetic architectures.

We used the R package “OMR” to implement the omnigenic MR analyses. The outputs for each OMR analysis include two primary parameters: the causal effect (α) and the proportion of SNP horizontal pleiotropy effect in the outcome variable (γ). Fig 1 displays the analysis framework of this study. Then We defined SCZ-triggering inflammatory markers as ones with a significant P value after Bonferroni multiple comparison correction (0.05/41 = 0.00122). Subsequently, these defined markers were chosen as the exposure in the second round of MR analysis of inflammation longitudinal morphology change of the brain, with the P value being corrected for Bonferroni multiple comparisons (0.05/15 = 0.00333).

### 2.3 Replication analysis

To assess the robustness of our main results, We carried out a two-layered replication analysis. For the first layer, we chose a different MR method, “MRMix”, a multiple-variant approach [26], to re-run the analysis on the inflammation markers showing a significant causal link with longitudinal brain changes in the previous step. For the second layer, we chose a different dataset, GWAS of multiple psychotic experiences”, to replicate the primary results of SCZ ⇒ inflammation [27].

## 3. Results

### 3.1. Seven inflammation markers showing significant change in response to schizophrenia

The results of SCZ⇒ inflammation analysis are summarized in Table 1 and Fig 2. In total, the omnigenic IVs of SCZ led to the change of seven inflammation markers in the peripheral blood. Of them, the most significant was CRP, with a causal estimate of −0.035 (α) in response to schizophrenia (P = 1.71 ×10^−32^); approximately 0.1% of SNPs involved indicative of a horizontal pleiotropy; the second most significant inflammation marker, B_NGF (basic nerve growth factor), decreased by 0.10 in the peripheral blood as the quantitative risk of SCZ increased each s.d unit. Further, We observed a systematic immunological deficits in the inflammatory response to SCZ, with only IL8 (interleukin 8) exhibiting an elevated level in response to schizophrenia α = 0.086, P = 3.64× 10^−7^). Of note, GROA (CXCL1), which could bind to the same receptor as IL8, CXCR2, was also significantly involved, with its mean value decreasing by 0.068 as the risk for SCZ increased each s.d unit, implying that a cytokine network including the IL8 pathway might be involved.As for the horizontal pleiotropy, most causal estimates of identified markers, except those of GROA (CXCL1) and MCSF, indicated varying yet relatively modest degrees of horizontal pleiotropy, signalling a potentially unremarkable associ no independent pathway other than through SCZ seems to be able to link SCZ to these two markers.

**Table 1:**
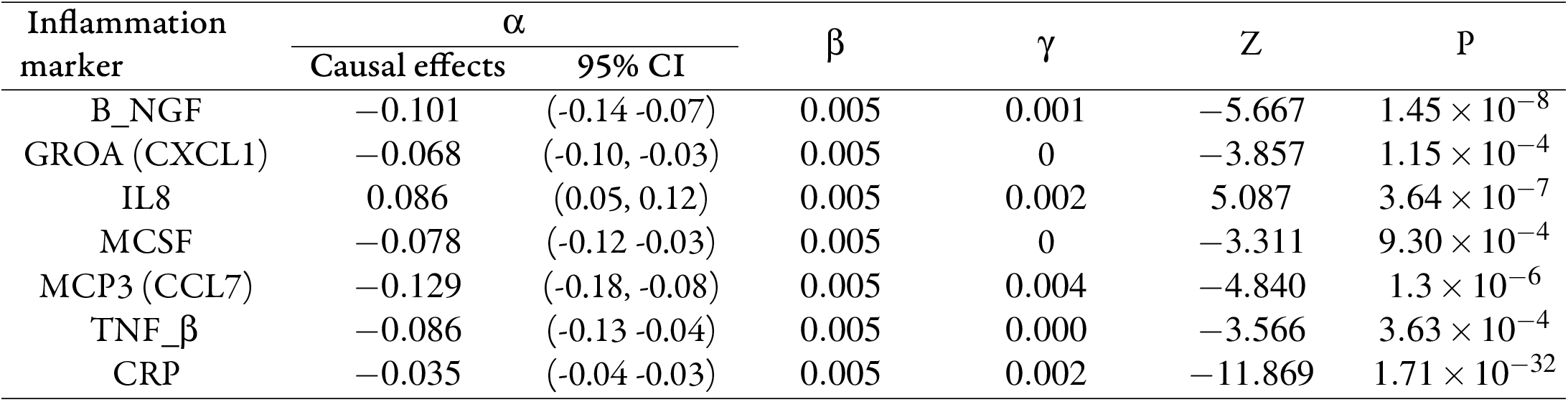
Causal estimates of six cytokines/chemokines and CRP in MR analysis of SCZ ⇒ inflammation

| Inflammation<br>marker | $\alpha$ | | $\beta$ | $\gamma$ | Z | P |
| --- | --- | --- | --- | --- | --- | --- |
|  | Causal effects | 95% CI |  |  |  |  |
| B_NGF | -0.101 | (-0.14 -0.07) | 0.005 | 0.001 | -5.667 | $1.45 \times 10^{-8}$ |
| GROA (CXCL1) | -0.068 | (-0.10, -0.03) | 0.005 | 0 | -3.857 | $1.15 \times 10^{-4}$ |
| IL8 | 0.086 | (0.05, 0.12) | 0.005 | 0.002 | 5.087 | $3.64 \times 10^{-7}$ |
| MCSF | -0.078 | (-0.12 -0.03) | 0.005 | 0 | -3.311 | $9.30 \times 10^{-4}$ |
| MCP3 (CCL7) | -0.129 | (-0.18, -0.08) | 0.005 | 0.004 | -4.840 | $1.3 \times 10^{-6}$ |
| TNF_$\beta$ | -0.086 | (-0.13 -0.04) | 0.005 | 0.000 | -3.566 | $3.63 \times 10^{-4}$ |
| CRP | -0.035 | (-0.04 -0.03) | 0.005 | 0.002 | -11.869 | $1.71 \times 10^{-32}$ |

**Fig. 2:**
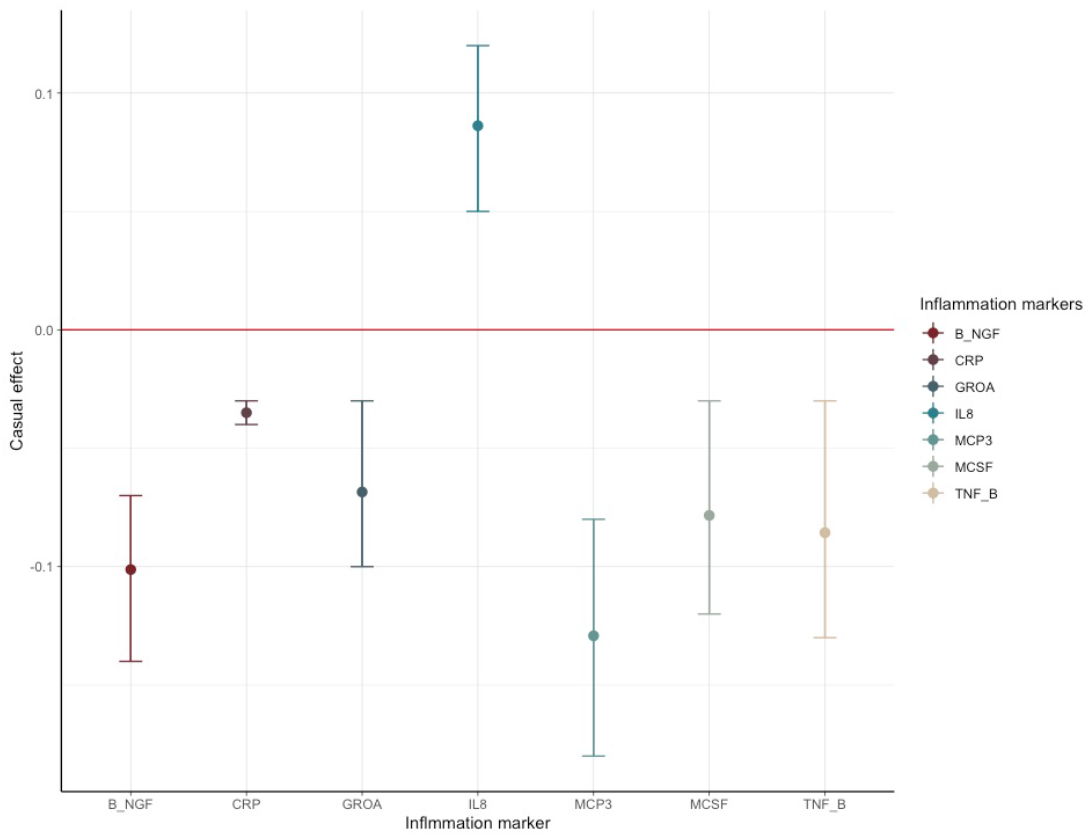
Causal estimates of seven inflammation markers responding significantly to schizophrenia

### 3.2. Three identified SCZ-triggering inflammation markers confering impact on the longitudinal morphology change of different brain regions

In the second round of MR analysis of inflammation⇒ the longitudinal brain changes, three out of seven SCZ-triggering inflammation markers, GROA (CXCL1), IL8 and CRP, caused the significant longitudinal change in the different brain regions.Specifically, CRP, out of three inflammation markers, as exposure, spawned the most significant and wide-ranging linear change in the brain morphology: one standard unit increase of CRP in the peripheral blood could lead to a decrease in the mean linear change rate by 0.03 in the white matter of cerebellum (P = 0.00067), an increase in the mean change rate of the cerebral white matter by 0.06 (P = 3.588× 10^−7^), a reduction in the change rate of pallidum by 0.05 (P = 4.02× 10^−8^), and an increase in change rate by 0.04 in the surface area (P = 8.67× 10^−5^). Besides, a nominally significant causal relationship could be established between GROA (CXCL1) and the linear change rate in lateralventricles (LCs), with a decrease in LCs mean value following an increased rate of 0.20 of GROA (P = 0.01). IL8, on the other hand, was casually associated with the linear change rate of cerebral white matter, with a causal effect estimated as 0.27 (P = 0.0064). The findings are summarized in the pathway diagram in Fig 3–5.

**Fig. 3:**
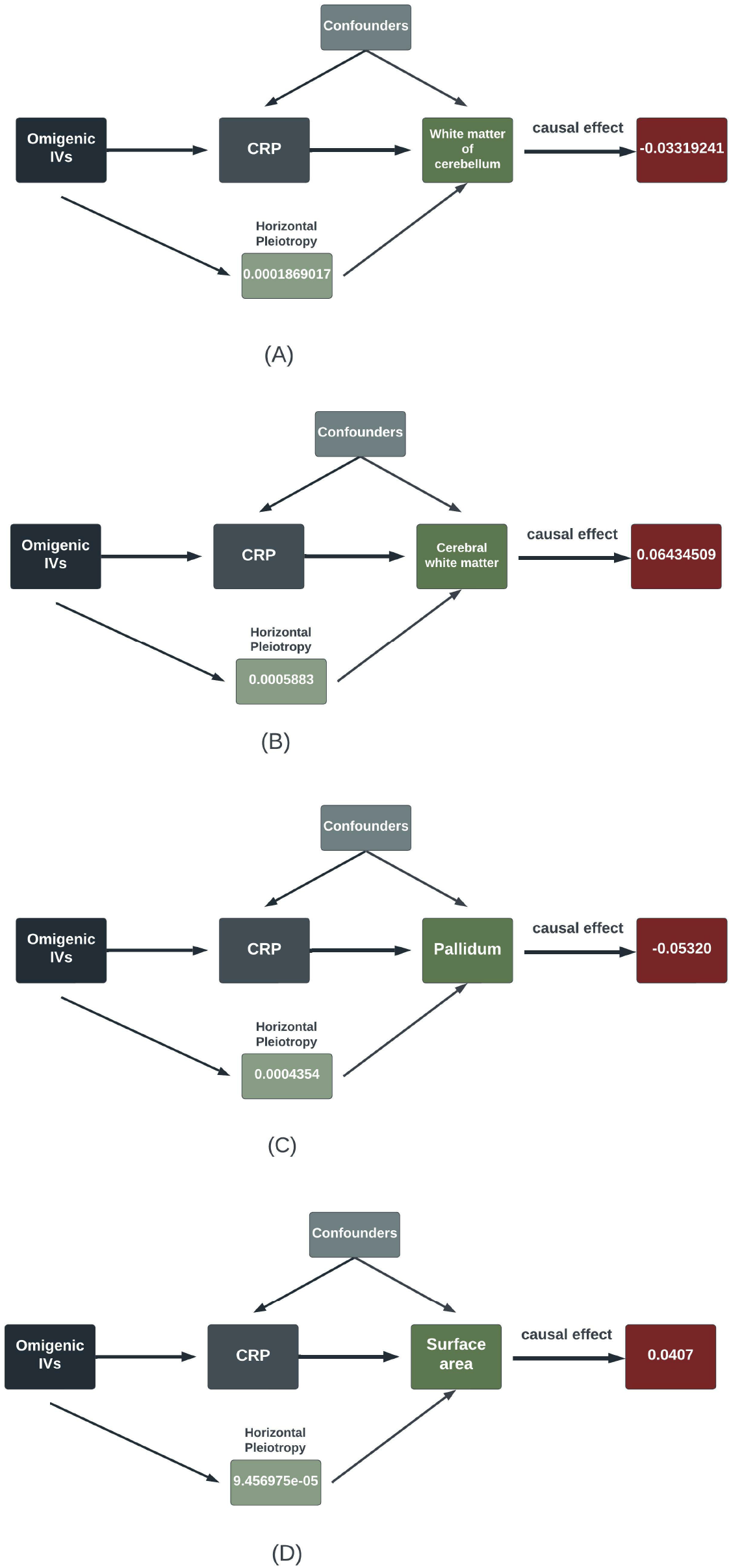
Estimates of the significant casual effect of CRP on the linear change rate of different brain regions (A) White matter of cerebellum (B) Cerebral white matter (C) Pallidum (D) Surface area

**Fig. 4:**
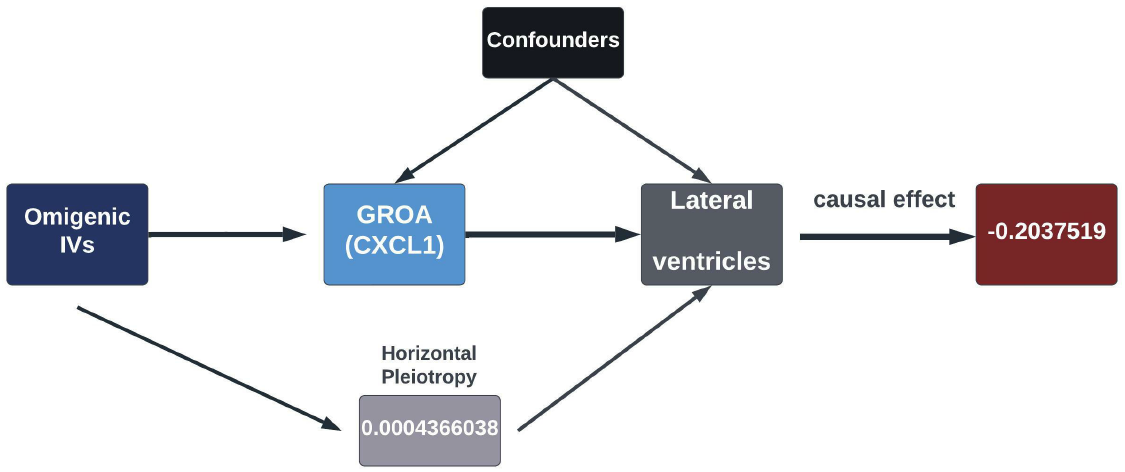
Estimates of the nominally significant casual effect of GROA (CXCl1) on the linear change rate of lateral ventricles

**Fig. 5:**
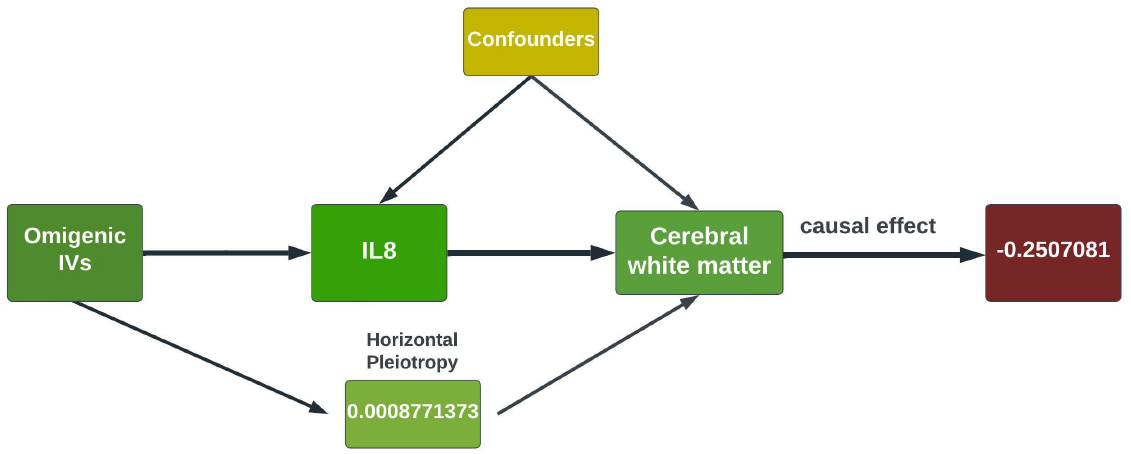
Estimates of the nominally significant casual effect of IL8 on the linear change rate of cerebral white matter

Replication using MRMix validated the significant causal relationship of schizophrenia to the three inflammation markers mentioned above. As displayed in Fig. 6, the direction of causal effects arising from MRMix was consistent with those from OMR for all markers except for GROA (CXCL1), which, instead of a negative causal effect, generated a positive estimate of 0.01. Using the GWAS summary statistics of multiple psychotic experiences from the UK biobank, we identified the causal relationship between multiple psychotic experiences and CRP to be nominally significant.

**Fig. 6:**
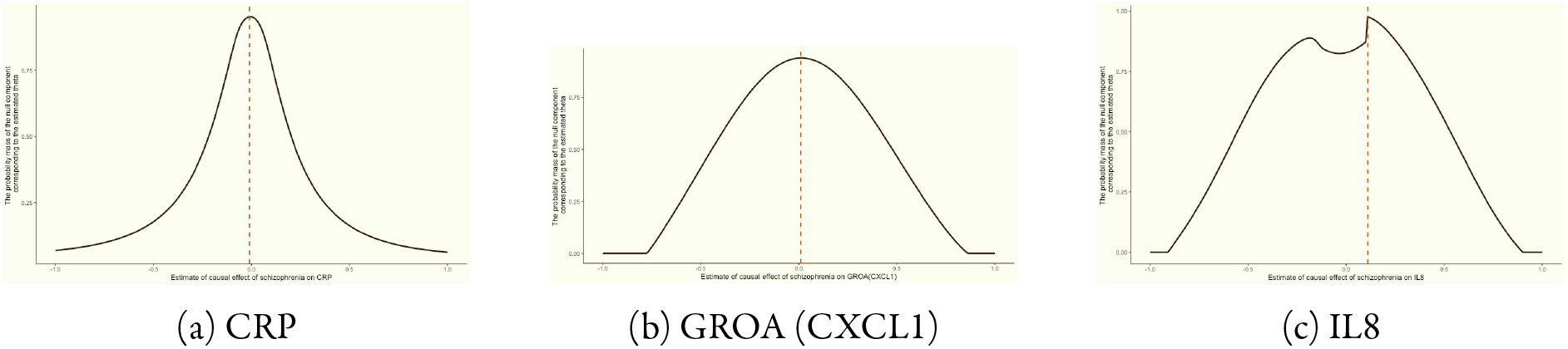
Grid search of possible causal effects (theta) of schizophrenia on CRP, GROA (CXCL1) and IL8; the maroon vertical line pointing to the theta which leads to maximum pi0

## 4. Discussion

For the first time, our current study used an omnigenic model to try to detangle the immune ramification of schizophrenia and its effect on the longitudinal changes of brain morphology. We identified multiple inflammatory markers linked to schizophrenia, suggestive of the systematic immunological deficits in schizophrenia. Also, we found three SCZ-triggering markers, CRP, GROA(CXCL) and IL8, could significantly affect the linear change rate of the brain, especially that of the white matter. The subsequent analysis using MRMix replicated our primary findings for all three markers, while using the result of “multiple psychotic experiences”, we detected the causal implication of CRP.

The primary findings arising in our current study are the causal relationship between schizophrenia and C-reactive proteins (CRP) and between the CRP and linear change rate of multiple brain regions. The mean value of CRP decreased by 0.035 as the quantitative risk of SCZ increased by one standard unit. CRP is an acute-phase protein mainly secreted by hepatocytes. In the central nervous system (CNS), CRP can be secreted by neurons in the face of infection [28]. CRP can exist either as a pentameric form (pCRP) or as separate monomers at the sites of inflammation (CRP). Our results recapitulate the previous findings that low-grade yet persistent CRP anomaly was intrinsic to the pathogenesis of SCZ and indicated that the pathogenetic factors of SCZ, such as intrauterine, stress, etc., lead to immunological deficits, exemplified by the decreases in the CRP level found in our study.

These immunological deficits, combined with environmental factors later in life, might contribute to the onset of schizophrenic symptoms. Meanwhile, our second round of MR analysis demonstrated that CRP could significantly give rise to deviant morphological changes in various brain regions, especially the white matter volume (WMV). It has long been recognized that inflammation could lead to white matter abnormality, such as lesions [29] and change in WMV [30]. Our results estimated an increase in the mean value of cerebral white matter and a decrease in the mean value of cerebellar WMV as CRP increases. A seminal study by Bethlehem et al., catalogued the morphological change of the brain over the human life span. In their study, white matter volume (WMV) increased rapidly from mid-gestation to early childhood, peaking at young adulthood, which overlapped with the onset age of schizophrenia [31]. Our results imply that the change in the CRP level caused by SCZ leads to a misaligned growth trajectory of WMV between cerebral and cerebellar areas. Our study, for the first time, established the causal link from CRP to the pallidum’s GMV changes which played a substantial role in various mental functions [32, 33]. Interestingly, Weiqiu et al., in their study, identified a ramarkable shared genetic architecture between schizophrenia and subcortical volumes, in particular, 54% for schizophrenia. Furthermore, this study also found that shared genetic loci reached the expression peak at the prenatal age, consistent with our assumption that the pathogenesis of schizophrenia, especially the one involving immunology, could start around the perinatal period, either triggered or precipitated by intrauterine infection or inflammation involving CRP [34].

Our study also identified another SCZ-triggering inflammation marker, GROA (CXCL1), that could lead to decreased linear change rate in the lateral ventricle. GROA is a protein structurally related to interleukin-8 (IL-8) and possesses potent neutrophil-stimulating activity [35]. Several studies have shown a significant difference in the gene expression and the DNA methylation between patients with SCZ and healthy controls and between the subtypes of schizophrenia [36]. In CNS, CXCL1 mainly binds to CXCR2 and promotes the proliferation of neuronal stem cells (NSCs) in both *vitro* and *vivo*, especially in the subventricular zone (SVZ) of the lateral ventricle [37]. Our analysis generated a negative causal estimate, meaning that SCZ could lead to a decreased level of GROA (CXCL1), which, in the long run, might suppress the proliferation and migration of NSCs.

Another SCZ-triggering inflammation marker which could confer impact on the linear change rate of brain morphology is interleukin 8, IL8. IL8 is a critical mediator of immune response [38]; in CNS, it is generally acknowledged that IL8 is synthesized by glial cells and astrocytes [39]. Our analysis of SCZ⇒ inflammation indicated a positive causal effect of schizophrenia on IL8,i.e. schizophrenia could result in an elevated level of IL8, which is consistent with many other studies [40, 41]. Moreover, we found that IL8 causes decreased change rate of cerebral white matter. Potential roles IL8 could play in neuroinflammation include facilitating the synthesis of pro-apoptotic protein, inducing the gene expression of proinflammatory proteases with neurotoxic properties, and recruitment of immune cells to the inflammation sites [8]. Of note, both IL8 and GROA (CXCL1) belong to the same CXC group, and they can bind to CXCR2 with high sensitivity [42]. Giovannelli et al., through patch clump technique and laser confocal microscopy, demonstrated that IL8 and GROA could modulate the Purkinje neuron activity in the mouse cerebellum [43]. Meanwhile, IL8 and GROA could cause significant longitudinal changes in cerebral white matter and lateral ventricle. One neuroimaging study detected a strong association between white matter diffusivity and ventricle volume in patients with Alzheimer’s disease (AD) [44]. A future study should be guaranteed to investigate the cellular mechanism of the IL8 pathway-mediated effect on the cerebral white matter.

Besides, we also identified B_NGF, MCSF, MCP3 and TNF_*beta*, the peripheral level of which could be altered by schizophrenia and which were not shown to be causally associated with the linear change rate of brain morphology. B_NGF is a neurotrophic factor that regulates functions of differentiated neurons [45]. Previous studies have found its dysfunction in schizophrenia, which correlates with a structural deviation of the brain in schizophrenia, including areas highly involved in crucial cognitive functions such as the prefrontal and temporal cortex [?]ciafre2020nerve). Our findings of a decreased level of B_NGF suggest a deficit neuro-developmental process preceded or precipitated by inflammation or infection. MCSF, Macrophage colony-stimulating factor, is a widely expressed cytokine. Its main functions include stimulating progenitor cells from bone marrow and subsequent development, proliferation and maintenance of mononuclear phagocytes such as dendritic cells and microglia [46]. MCSF, in the presence of anti-inflammation factors such as TNF_*beta*, is capable of polarizing microglia towards anti-inflammatory (M2) directions [47]. Both MCSF and TNF_*beta* showed lower levels in response to the pathogenesis of schizophrenia, hinting at a possibly early-onset dysfunction in restricting neuroinflammation. MCP3 (CCL7), the monocyte chemotactic protein 3, belongs to the MCP subgroup of CXC chemokines and is a potent chemoattractant for a variety of leukocytes including dendritic cells (DCs) [48]. Cathomas et al. compared a series of cytokines between patients with schizophrenia and healthy controls and investigated the relationship between cytokines and quantitative traits of SCZ; their results, consistent with our estimate, indicated a significantly lowered level of CCL7 in patients with SCZ [49]. Our study did not detect any causal relationship between the five cytokines herein and the linear change in brain morphology. It could be speculated that they might confer their risk through other pathophysiological processes, possibly in a time-specific manner.

## 5. Limitations

To the best of our knowledge, this is the first study to use an omnigenic conceptual framework to establish the inflammatory implications of schizophrenia and the effect of these inflammatory effectors on the temporal change of brain morphology. Although we ensured the robustness of our results by replicating preliminary results using a different method and dataset, we are fully aware of the limitations of our present study. First, we can not exclude all possible confounders in our MR analysis, violating the exclusion restriction assumption. Due to the polygenicity, or even omnigenicity of complex diseases and traits, widespread pleiotropy exists in the human genome, and it is currently implausible to identify all potential confounding factors. Secondly, we only took a uni-directional MR analysis, not exploring the causality from cytokine to schizophrenia out of concern that IVs of cytokines would be largely biased toward underpowering due to the modest sample size of cytokine GWAS.

## 6. Conclusion

In summary, we used an omnigenic conceptual framework to identify the inflammatory effectors of schizophrenia and the effect of these effectors on the temporal variation of brain morphology. The results identified systematic, albeit low-grade, immune modifications triggered by the pathogenesis of schizophrenia. Further, these immune modifications could temporally confer their risk of schizophrenia by disrupting the structural integrity in both cortical and subcortical areas.

## Data Availability

Summary statistics of SCZ GWAS: https://doi.org/10.6084/m9.figshare.19426775;
Summary statistics of GWAS of cytokine level in the peripheral blood: https://research-information.bris.ac.uk/en/datasets/cytokines-gwas-results;
Summary statistics of GWAS of longitudinal change of brain morphology: http://enigma.ini.usc.edu/research/download-enigma-gwas-results

